# Consultation rate and mode in English general practice, 2018 to 2022: a population-based study by deprivation

**DOI:** 10.1101/2022.12.06.22283150

**Authors:** Emma Vestesson, Kaat De Corte, Elizabeth Crellin, Jean Ledger, Minal Bakhai, Geraldine M. Clarke

## Abstract

**Background:** The COVID-19 pandemic has had a significant impact on primary care service delivery. With general practice delivering record numbers of appointments and rising concerns around access, funding and staffing in the UK National Health Service, we assessed contemporary trends in consultation rate and mode (face-to-face versus remote).

**Methods:** We did a retrospective analysis of 9,429,919 consultations by GP, nurse or other health care professional between March 2018 and February 2022 for patients registered at 397 English general practices. We used routine electronic health records from Clinical Practice Research Datalink Aurum with linkage to national datasets. Negative binomial models were used to predict consultation rates and modes (remote versus face-to-face) by age, sex, and socio-economic deprivation.

**Findings:** Overall consultation rates increased by 15% from 4.92 in 2018-19 to 5.66 in 2021-22 with some fluctuation during the start of the pandemic. Consultation rates increased with deprivation. The breakdown into face-to-face and remote consultations shows that the pandemic precipitated a rapid increase in remote consultations across all groups but varies by age. Socioeconomic differences in consultation rate, adjusted for sex and age, halved during the pandemic (from 0.36 to 0.18 more consultations in the most deprived). The most deprived saw a relatively larger increase in remote and decrease in face-to-face consultations rates.

**Interpretation:** Substantial increases in consultation rates imply increased pressure on general practice. The narrowing of consultation rates between deprivation quintiles is cause for concern, given ample evidence that health needs are greater in more deprived areas.

**Funding:** No external funding.

**Research in context:** *Evidence before this study:* Pressure on general practice has increased over recent years and there is consensus that the COVID-19 pandemic added to this. There is also consensus that the way general practice is delivering care has changed with increased use of remote consultation but there no estimates of the full extent of this and uptake by different groups. A seminal paper - Clinical workload in UK primary care: a retrospective analysis of 100 million consultations in England, 2007–14 – found an increase in consultation rates over the study period and in increased reliance on telephone contacts even before the pandemic.

*Added value of this study:* This study reports recent data for general practice consultation rates overall and by delivery mode. Our findings show that overall consultation rates were higher in 2021-22 than prior to the pandemic and that there has been a shift from face-to-face to remote consultations. However, the increase in overall consultations rates varies between index of multiple deprivation quintiles when adjusting for age and sex. These findings are based on close to 10 million consultations and 2 million person-years of observation from a validated data base of routinely collected electronic clinical records (the Clinical Practice Research Datalink Aurum).

*Implications of all the available evidence:* Our analysis shows that general practice is busier than ever. We provide details on the use of remote versus face-to-face consultations by different patient groups over time. The narrowing of the difference between consultation rates of deprivation quintiles implies increasing health inequality in the population as existing differences in health needs are therefore not fully reflected in the consultation rates. The relatively larger increase in remote consultation rates and drop in face-to-face consultations for the most deprived provides detail on what type of consultations different patient groups receive but also raises additional questions.

## Introduction

In the last few years, digital technology has enabled new ways of working in general practice. Before the COVID-19 pandemic, remote consultations (by telephone, video, text based and online) were steadily increasing on a background of supporting policy. The NHS Long Term plan 2019, committed to every patient having the right to digital-first primary care by 2023/24; the 5-year GP contract reform framework, committed for all practices to offer online consultation systems by 2021. The COVID-19 pandemic served as a catalyst for the uptake of remote consultations as guidance was issued to triage patient contacts wherever possible encouraging the use of remote consultations where clinically appropriate to avoid risks of COVID exposure to patients and staff. (1) Before the pandemic it was reported that less than 30% of consultations were carried out remotely; within weeks it was 77%. (2)

Both remote and face-to-face consultation modes have advantages and disadvantages. Remote consultations can offer potential benefits through expanding access to services and appointment flexibility – in particular, for patients in rural areas, patients who find face-to-face consultations difficult and those with substantial barriers for travelling to their general practice such as mobility issues, employment or caring commitments. Faster access to care and a more cost-effective alternative to face-to-face appointments have also been highlighted as potential benefits.(3) In this way, remote consultations could decrease some of the current inequalities observed in the usage of primary care.

On the other hand, there are concerns that remote consultations could exclude certain patient groups and exacerbate health inequalities. (4) Though the evidence around remote consultations and inequalities is limited, there is ample evidence that primary care is under more pressure in more deprived areas so understanding the impact of a rapid increase in remote consultations is important. (5) Factors such as age, disability, digital exclusion, communication needs, data poverty and lack of trust can impact people’s willingness or ability to access and benefit from remote consultations. A systematic review conducted at the start of the pandemic collated evidence on remote versus face-to-face consultations with a focus on inequalities and observed variation in the use of remote consultations, but the evidence was not conclusive. (6) A study of remote primary care for people experiencing homelessness during the pandemic found that the shift to remote consultation created barriers to accessing care due to factors such as the lack of funds to make calls or access to a telephone. (7) A cross-sectional study observed that there are differences in the proportion of consultations delivered remotely by category of the index of multiple deprivation (IMD). (8) However, a longitudinal study did not observe a difference in the change of proportion of remote consultation over time by deprivation. (3) Many of these studies are from before or the inception of the pandemic and the use of remote consultations has substantially changed. Studies from outside the UK do observe inequalities but due to the difference in health systems, these results are unlikely to generalise to the English population.

In this study we use person-level data from a large, nationally representative sample to describe contemporary patterns in consultation rates and modes in English general practice before and during the COVID-19 pandemic. We further investigate the inequalities in consultation rates and modes between individuals grouped by age, sex, and deprivation.

## Methods

### Study design and data

We performed a cohort study using person-level data from the Clinical Practice Research Datalink (CPRD) Aurum between January 2018 and February 2022. CPRD Aurum is a database with routinely collected data from general practices in England that use the EMIS Web® information management system. CPRD contains data for over 40 million patients from 1,332 practices in England as of May 2022. (9) Patients are broadly representative of the English population based on age, sex and deprivation. CPRD provided linkage to the 2015 index of multiple deprivation (IMD) at the patient level based on the patient’s LSOA and to the 2011 Urban-Rural classification based on the practice LSOA. The study protocol was approved by CPRD’s Research Data Governance (protocol number: 21_000357).

### Eligibility criteria

We studied patients registered at general practices that participated in CPRD Aurum. Eligibility criteria were applied at both practice and patient level. 400 practices located in England were sampled at random. Eligible patients were those with acceptable data quality (verified by CPRD); registered at one of the 400 practices at any point between January 2018 and May 2021; recorded as either male or female sex; and eligible for area level linkage to the IMD 2015. From this cohort, a sample of 600,000 patients was randomly sampled. Three GP practices were identified by CPRD as having duplication issues and therefore excluded. In addition, two patients were excluded as they no longer met the inclusion criteria after the final dataset was extracted.

### Consultation type and consultation mode

The primary information source on consultations was the CPRD Aurum ‘Consultation’ table which contains information on the type of consultation (e.g., telephone, home visit, practice visit). The consultation table can be linked to the ‘Staff’ table to gain information about the staff member and the observations that occurred during the consultation. We included consultations carried out by GPs, nurses and other general practice care providers. Clinical consultations were identified. Consultations that were not attended were excluded. This builds on methods used on CPRD Aurum. (10)

Consultations were further classified by mode of delivery as either remote (by telephone, video or SMS/online) or face-to-face (at the surgery or at home) consultations based on information in the consultation table for consultation mode or observations recorded during the consultation (Table S1). Where a consultation’s mode was unclear, it was assumed to be face-to-face. A patient could have multiple consultations per day with a mix of modes. The final dataset included consultations for patients between March 2018 and February 2022. For year-on-year comparisons, we grouped consultations in 12-month periods (March 2018 - February 2019, March 2019 - February 2020, March 2020 - February 2021 and March 2021 - February 2022), covering two years pre-pandemic and two years during the pandemic.

### Statistical analysis

We calculated person-years of observation over the study period based on the patient’s registration dates and when the practices submitted data to CPRD. Crude consultation rates and the proportion of remote consultations were calculated for each month and 12-month periods. To compare consultation rates over time, the number of consultations, remote consultations and face-to-face consultations per patient were modelled using a negative binomial regression with person-years as an offset. The models included age, sex, deprivation, and pandemic year as well as interactions between these terms. A small number of patients without a deprivation quintile were excluded from the models.

From these models, we estimated adjusted consultation rates for the 12-month periods for patients grouped by deprivation quintile, age, and sex to allow absolute estimates of the differences between these groups. These rates were calculated for the ‘average’ patient with respect to all the other variables in the model.

All analyses were carried out on a secure analysis server at the Health Foundation using R4.0.3, with the ggeffects package for estimating predicted values.

## Results

### Consultation rate

The study included 9,429,919 consultations over 1,863,507 person-years of observation from 397 practices located across all regions in England between March 2018 and February 2022. Overall, face-to-face and remote consultation rates did not change materially between 2018-19 and 2019-20 (Table 1, Figure 1). Consultations averaged 4·92 and 4·94 per person-year in 2018-19 and 2019-20 respectively with approximately 4 times as many face-to-face (4.00 and 4.02 per person-year in 2018-19 and 2018-20 respectively) as remote (0·91 and 0·92 per person-year in 2018-19 and 2018-20 respectively) consultations. During the pandemic, consultation rates initially dropped from 4·92 per person-year in 2018-19 to 4·76 per person-year in 2020-21, but by 2021-22 had increased to 5·66 per person-year, 15% higher than 2018-19 (Table 1, Figure 1).

**Table 1.** Temporal trends in GP, nurse and other HCP consultations by subtype of face-to-face and remote consultation mode. Crude rates (per person-year)

|  | 2018-19 | 2019-20 | 2020-21 | 2021-22 | Change between 2018-19 and 2021-22 |
| --- | --- | --- | --- | --- | --- |
| All GP, nurse and other HCP consultations per person-year (%) |  |  |  |  |  |
| All modes | 4.92 | 4.94 | 4.76 | 5.66 | 15.0% |
| Face-to-face | 4.00 (81.3) | 4.02 (81.4) | 2.32 (48.7) | 3.32 (58.7) | -17.1% |
| Remote | 0.91 (18.5) | 0.92 (18.6) | 2.45 (51.5) | 2.34 (41.3) | 155.3% |

**Figure 1.**
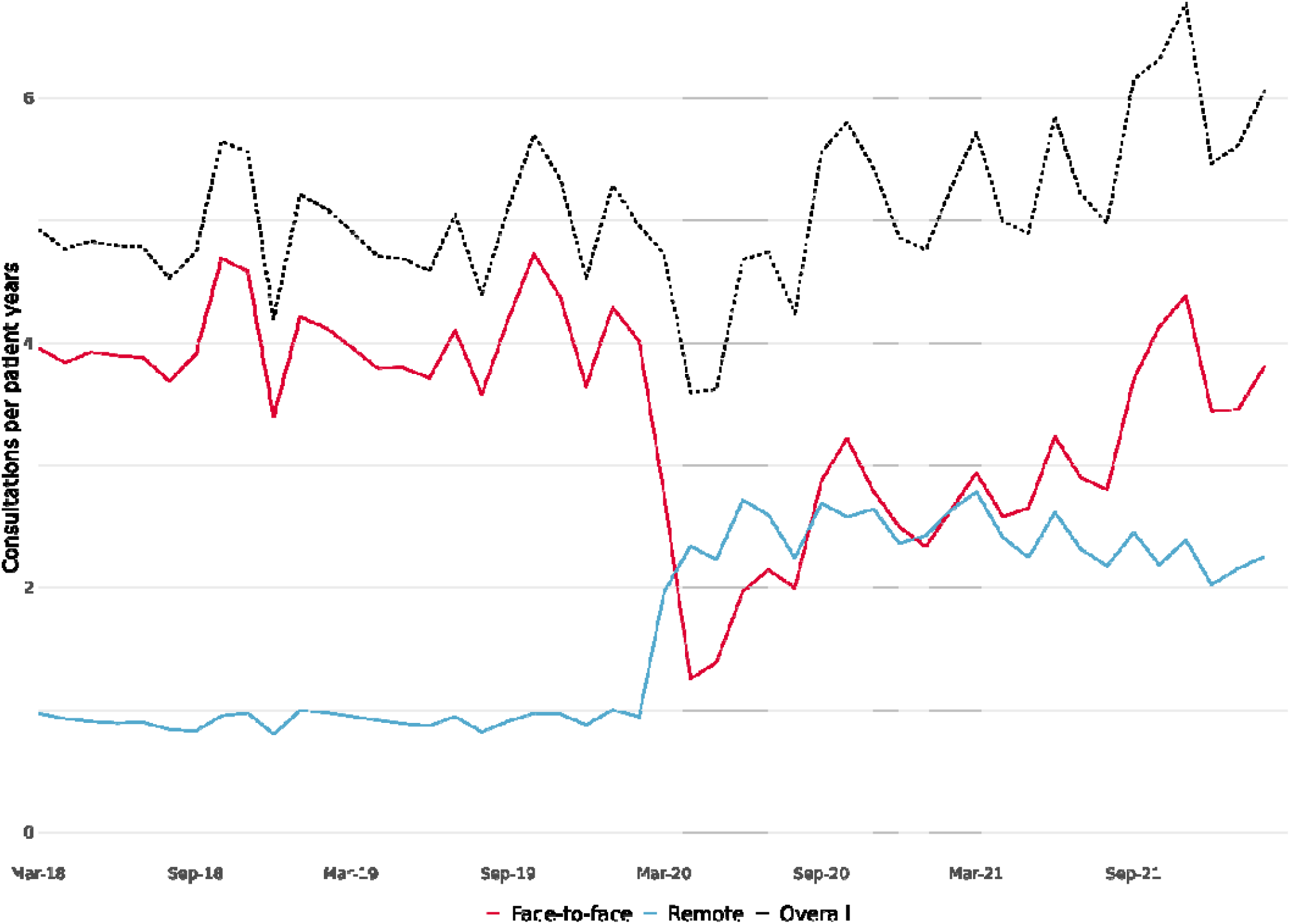
Overall consultation rates and consultation rates by delivery mode

### Consultation mode

Over our study period there were dramatic shifts in consultation mode. In 2020-21, compared with 2018-19, the rate of face-to-face consultations almost halved from 4·00 per person-year to 2·34 per person-year while the rate of remote consultations more than doubled from 0·91 to 2·45 per person-year. During 2021-22, rates of face-to-face consultation steadily recovered while the rate of remote consultations remained stable, and by the end of our study period in February 2022, more consultations were delivered face-to-face (63%) than remotely (37%) (Supplementary Figure 1). In the last year of our study 2021-22, rates of face-to-face consultation were 17·1% lower and rates of remote consultation were 155·3% higher, compared with pre-pandemic 2018-19.

**Table 2.**
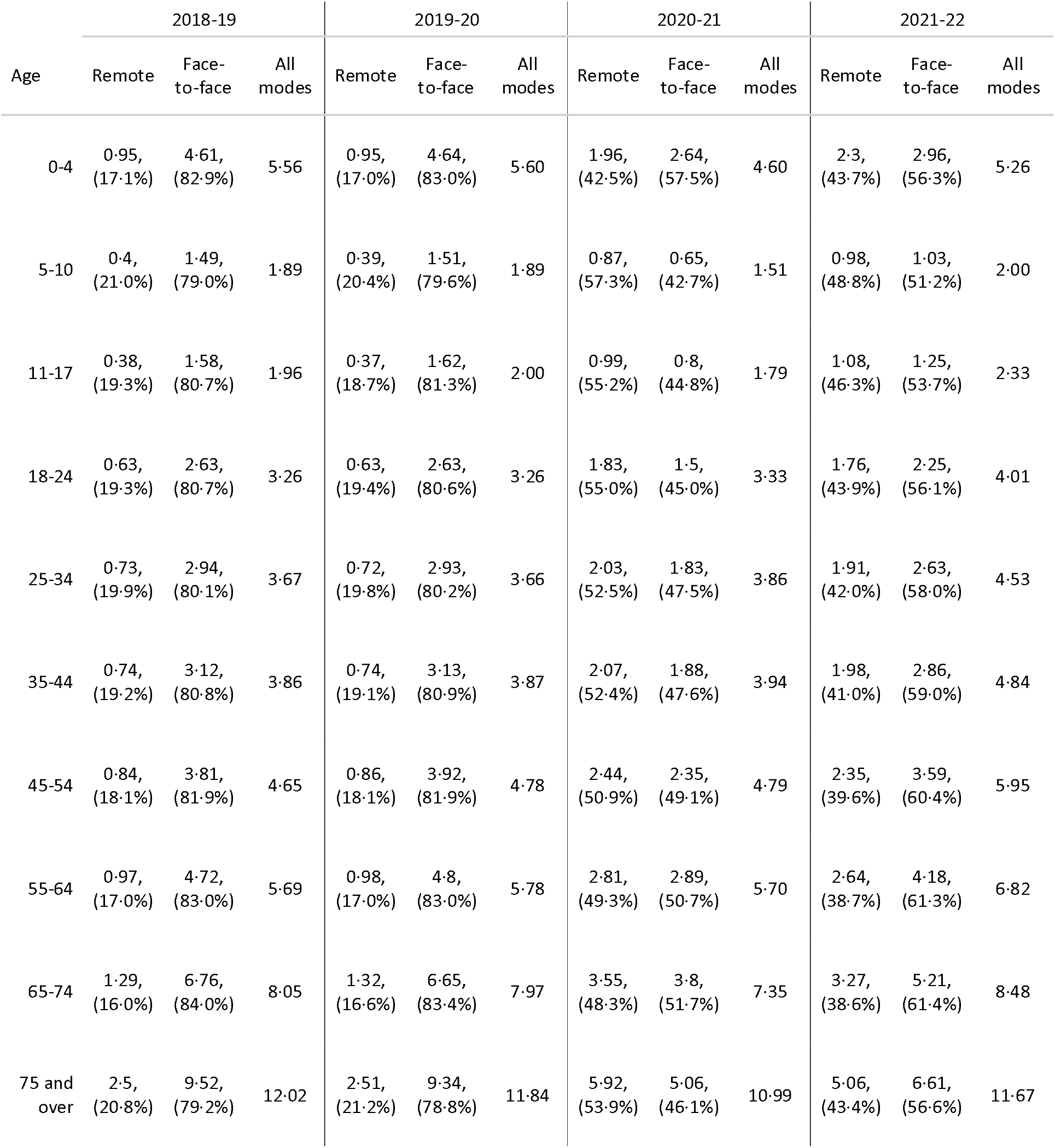
Consultations by mode and age Consultation rates per patient years and percent of all consultations within age group and pandemic year

### Variation by age

Age-specific consultation rates had a consistently J-shaped distribution across all years with the highest rates in the youngest patients aged 0-4 years, decreasing to lowest levels in children aged 5-10 years before steadily increasing in each age group to a peak in patients aged 75 years and over.

Between 2018-20, consultation rates remained fairly steady within each age group with little variation in the proportion of consultations delivered remotely. In 2020-21, a year-on-year decrease in the overall consultation rate was driven by those in the youngest (aged 0-17 years) and oldest (aged over 55 years) age groups. The decline was most pronounced in infants (aged 0-4 years) although this group sustained the highest proportion of face-to-face consultations (57.5%) during this first year.

In 2021-22, consultation rates recovered to higher than pre-pandemic levels for all patients except those in the youngest (aged 0-4 years) and oldest (aged over 75 years) age groups. There was markedly less variation in consultation mode across the age groups in 2021-22 compared with 2020-21.

### Variation by deprivation

From the multivariable analysis, we present predicted age and sex-adjusted consultation rates by deprivation quintiles over time (Figure 3 and table S2). Each quintile represents 20% of local areas in England.

**Figure 2.**
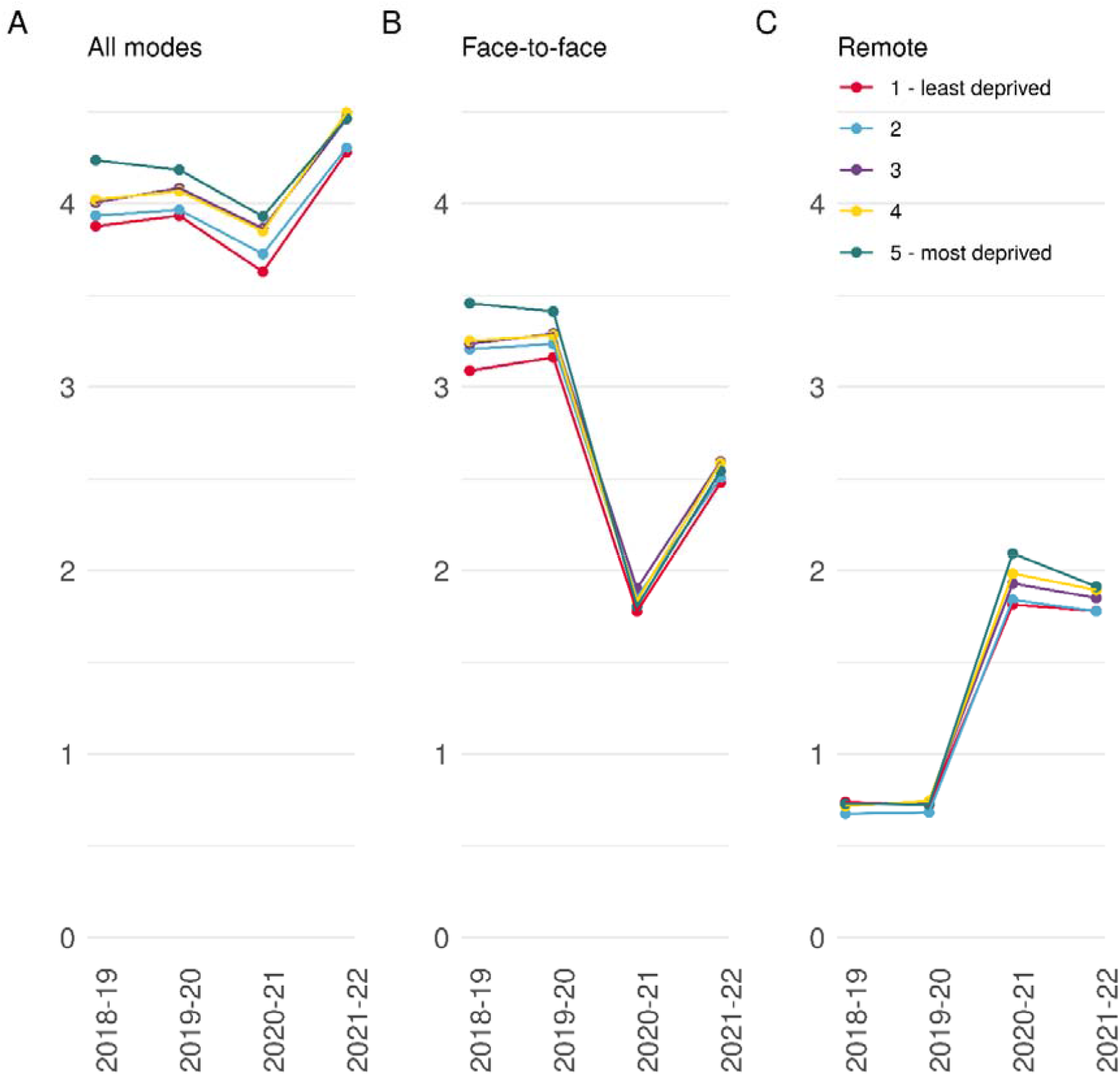
Consultation rates stratified by deprivation quintiles (A) Overall consultation rate per person per year (B) face-to-face consultation rate per person per year (C) remote consultation rate per person per year

**Figure 3.**
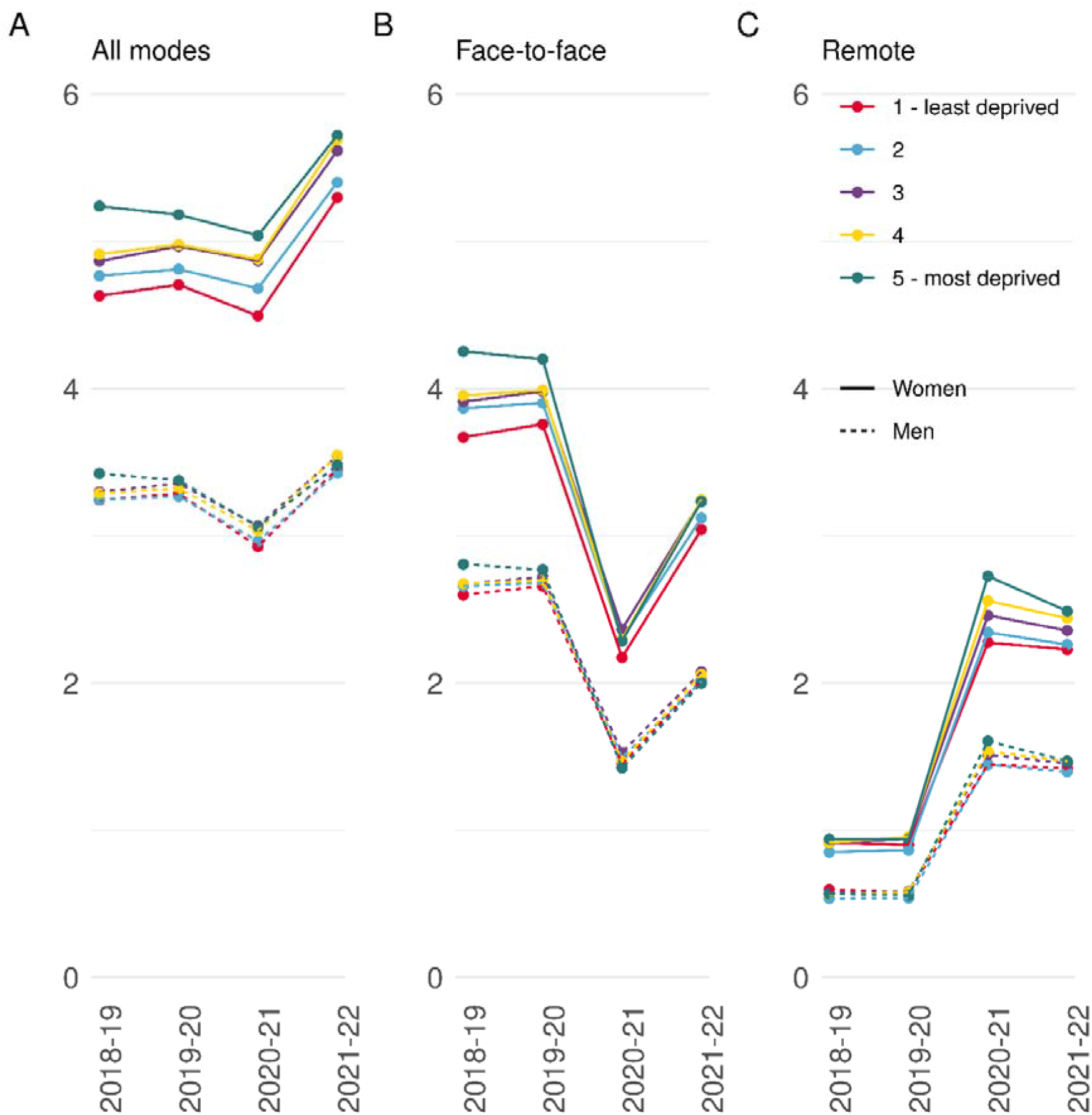
Predicted consultation rates over time by sex (A) Overall consultations per year by deprivation (B) face-to-face consultations per year by deprivation (C) remote consultations per year by deprivation

Consultation rates increased with increasing deprivation in each study period and were at their highest in the second year of the pandemic (2021-22) for all deprivation quintiles (Figure 3A). Over the study period consultation rates increased by 10% in the least deprived quintile, compared with 5% in the most deprived quintile, effectively narrowing relative differences between deprivation quintiles.

Rates of face-to-face consultation also tracked deprivation quintiles with patients living in higher deprivation quintiles consistently having a greater rate of face-to-face consultations than those living in lower deprivation quintiles (Figure 3B). Inequalities in rates of face-to-face consultation also narrowed during the pandemic with differences between the most and least deprived quintiles decreasing from 0·37 per person-year in 2018-19 to 0·06 per person-year in 2021-22.

There was little difference in the rate of remote consultation across deprivation quintiles before the pandemic (Figure 3C). During the pandemic, inequalities in rates of remote consultation widened, and patients in the higher deprivation quintiles had consistently higher rates than those in lower deprivation quintiles. In 2021-22, remote consultation rates ranged from 1·91 per person-year in the most deprived quintile to 1·78 per person-year in the least deprived quintile.

Figure 4 presents consultation rates by deprivation quintile and sex over time (see also Table S3). Notably, men have consistently lower consultation rates than women. Trends in the rate of face-to-face and remote consultation rates across deprivation quintiles for women and men are similar to that observed for the overall population except that differences between the quintiles are narrower in men compared with women. For example, in 2018-19 the difference in the consultation rate for men in the most compared with the least deprived quintile was 0·18 per person-year whereas for women it was 0·61 per person-year.

## Discussion

We provide contemporary, nationally representative data on the rate and mode of clinical consultations by GP, nurse, and other HCP staff in general practice, analysing trends across patient age, sex and deprivation over 1 million patient-years. These are the first comprehensive data to break down consultations by mode. Consultations rates in primary care are higher in 2021-22 than before the pandemic and the pandemic has led to a dramatic reconfiguration of consultation modes in English general practice with substantial heterogeneity across patient age, sex and deprivation.

The consultation rates calculated in this analysis are lower than those observed by Hobbs et al. (11) between 2007-2014. However, they are consistent with national data and due to variations in methodology direct comparisons can be difficult. Nevertheless, patterns across age and sex are consistent with those reported by Hobbs et al.

Before the pandemic more than 75% of consultations were face-to-face. After a sharp drop at the start of the pandemic, the proportion of face-to-face consultations has slowly increased but remains below pre-pandemic levels at the end of our study. This reflects a significant shift in general practice processes with the adoption of a hybrid approach with a mix of appointment types. These trends in consultation mode have been observed elsewhere but not in a large nationally representative sample. (3,6,7,12,13)

The dip in the proportion of face-to-face consultations at the start of the pandemic likely reflects operational guidance at the time. The pandemic created a driver for change. (14) Before the pandemic the implementation of remote consultations was slow due to a combination of technological difficulties, confidence and concerns about quality of care and low uptake from patients. (15) The model of general practice changed as a result with more practices using doctor-led remote triage as the access point for services. (2) The subsequent increase in face-to-face consultations, despite repeated waves of high COVID-19 infection rates, likely reflects the embedding of processes to manage infection control risk, a better understanding of the risks of the virus as well as changing government guidance on lockdowns and restrictions, although it is expected levels will remain below pre-pandemic levels to reflect new ways of working.

In the first year of the pandemic, the overall drop in the year-on-year consultation rate was driven by children (aged 0-17 years) and older patients (aged over 65 years). COVID restrictions such as home schooling, social distancing and shielding recommendations reduced the spread of some diseases especially non-COVID respiratory infections which may be reflected in the lower consultation rates. (16) On the other hand, some age groups saw an increase in consultation rates in the first year of the pandemic which could be COVID related consultations. Despite the drop in overall consultations for these groups, they sustained higher proportions of face-to-face consultations. This reflects an active prioritisation of groups by health need as they are more likely to present with issues that are more complex or difficult to assess remotely.

We observed consistently higher consultation rates in patients living in more deprived compared with less deprived areas. This tallies with evidence that patients living in more deprived areas are more likely to have higher health needs. (17) The pandemic likely had a negative impact on everybody’s health, this is reflected in all quintiles of deprivation experiencing higher consultation rates in 2021-22 than in the pre-pandemic years. However, over the course of the pandemic, differences between the most and least deprived quintiles reduced by five percentage points, reflecting overall larger increases in consultation rates for patients living in the least deprived populations. During the pandemic we know that the health of patients in more deprived areas worsened faster compared with those patients in less deprived areas. Therefore, it seems unlikely that inequalities in health care needs have decreased over the pandemic.

Moreover, our findings may indicate that the demand-capacity gap has widened for patients living in more deprived compared to less deprived areas. Previous research highlighted that practices in more deprived areas manage 10% more need adjusted for population size and receive 7% less funding adjusted for need compared with practice in more affluent areas. (18) In addition, they face greater workforce challenges with lower staff to patient ratios, recruitment rates and more staff absences due to illness during the pandemic. (19) Overall, the evidence points to existing significant pressure limiting the capacity to stretch services further in response to pandemic driven demand exacerbating health inequalities. This indicates a need for greater support and investment in services in deprived areas.

At a patient level, reasons why patients in deprived areas had relatively fewer consultations during the pandemic may include a higher risk of being infected or getting severely ill with COVID-19 (20), a greater likelihood of lower levels of health and digital literacy (21) and an uneven economic impact (22). These factors may have combined to result in both higher anxiety levels and greater concerns around contacting healthcare services due to risk of exposure to infection alongside greater difficulty in taking time off work to attend to healthcare needs. These findings correlate with reported patient overall experience of making an appointment where the fall in satisfaction rates was greater for practices in more deprived areas. (23) A greater reliance on technology could have also been a barrier to access for some patients in more deprived areas. (24)

During the pandemic the increase in remote consultation was reflected across all deprivation quintiles but was relatively larger for those living in the most deprived quintile - the difference in consultation rate between the most and least deprived populations increased by 13 percentage points for remote consultations in contrast to a decrease of 10 percentage points for face-to-face consultations. The reasons for the relatively greater use of remote consultations in more deprived populations during the pandemic are likely many and complex relating to both practice and patient level effects. At the practice level, staff at practices in more deprived areas are more likely to be older or from ethnic minority backgrounds known to have been more impacted by COVID-19. (20,25) Consequently, more staff might have been working from home in more deprived practices with more care being delivered remotely. At the patient level, socioeconomic variation in need could have played a role, as patients in more deprived quintiles are more likely to have long-term conditions and multimorbidity (26). Patients living in deprived areas are both at higher risk of being infected and getting severely ill with COVID-19 which could lead to more remote consultations. Individual circumstances such as ability to travel to the practice could also impact the choice of modality. (27)

Women having higher consultation rates than men has been widely reported (11). Interestingly, sex differences in consulting rates varied by deprivation status, reflecting a greater socioeconomic gradient in consulting rates among women than men. This finding has been observed elsewhere (28) and could be a consequence of differences in the number of comorbidities between women and men which also increases with increasing deprivation. (29)

Digital technology will play an increasing role in general practice, and a deeper understanding of what mode of consultation suits particular patients’ needs and preferences is continuing to develop. There is limited evidence on differences in clinical outcomes and patient satisfaction between remote and face-to-face consultations. However, a recent systematic review suggests that overall system design including the triage process and technology interface is key to good outcomes especially staff workload, but more research is needed on a broad range of outcomes. (30) Most remote consultations in England were telephone based and easier to access than other more technology heavy remote consultation options such as video calls. Despite the potential of digital tools to improve service delivery, it is vital to continuously improve their accessibility and usability and monitor the impact on health inequalities.

This study has several strengths. Using a rich clinical nationally representative patient-level dataset that includes more than 1 million patient-years allows us to explore consultation mode by different patient groups. Comparisons between deprivation quintiles were adjusted for age and sex making comparisons more accurate. We developed a novel method to derive consultation mode that combines both information from the time of booking with what happened during the consultation to improve the accuracy of our results.

Data quality is one of the main limitations of this study as the consultation mode is not consistently recorded in the data. The consultation mode was derived using a new method that has not been externally validated. The default assumption was that the consultation was face-to-face, we are therefore more likely to have underestimated the number of remote consultations. However, we found a strong increase in the use of remote consultations during the pandemic, with a peak around 75% of consultations being remote, which is consistent with other sources giving us confidence in our methods. (13)(14)

In summary, we find that general practice is delivering more consultations than ever but that trends in consultation rates and modes across deprivation quintiles indicate exacerbation of existing socioeconomic inequalities. This general increase in the rate of consultations matches other evidence and reports from GPs but further research is needed to ensure that consultation rates match health needs across deprivation groups.

## Data Availability

Data is available from CPRD but restrictions apply to the availability of this data which was used under license for the current study.

## Supplementary materials

*Table S 1*

**Figure S 1.**
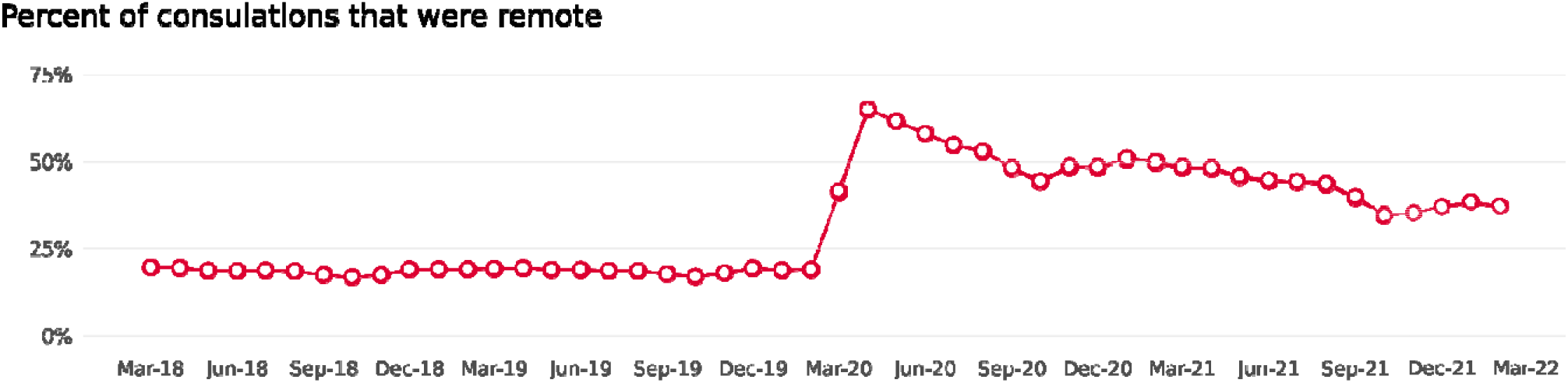

**Figure S2.**
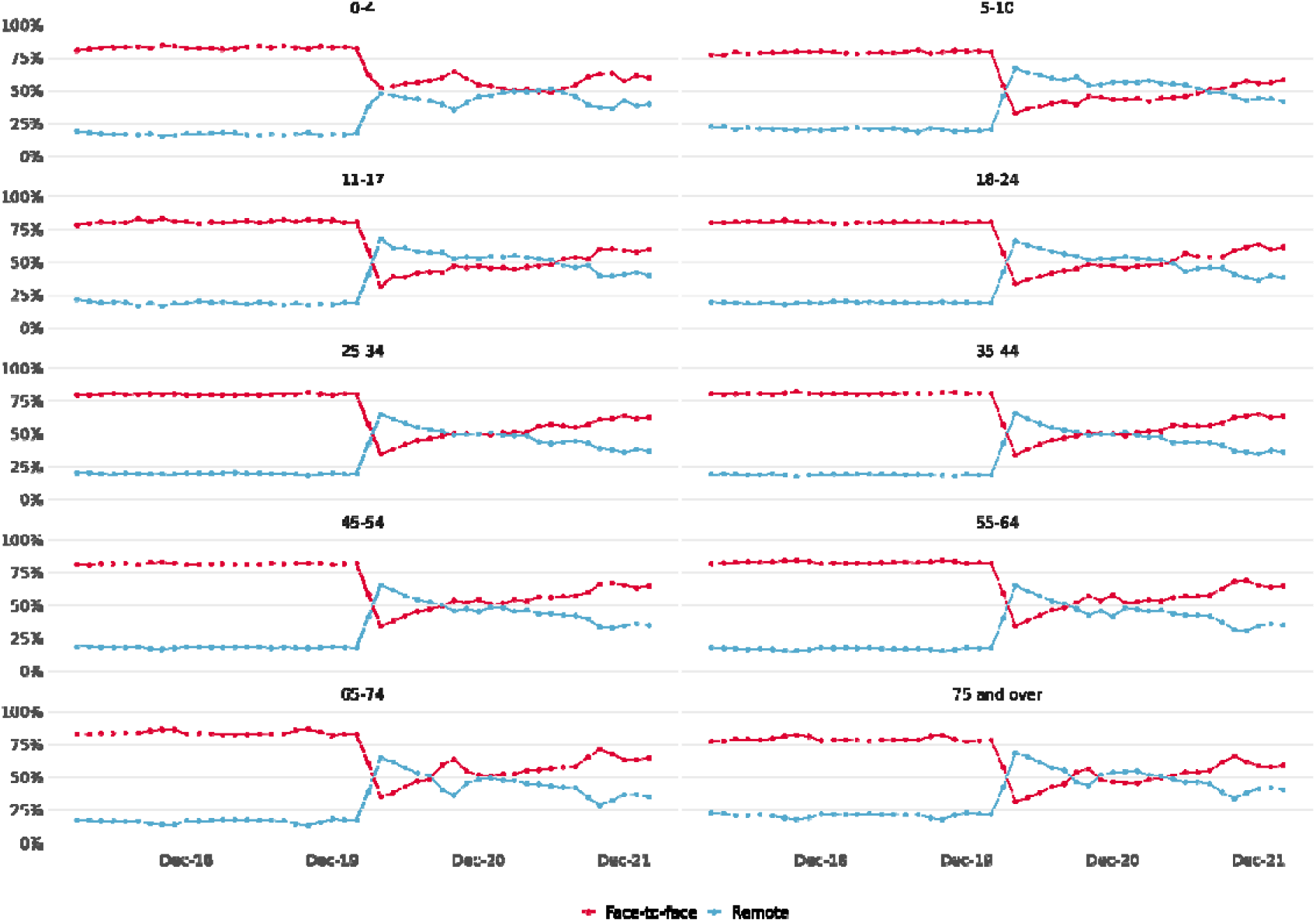
Percentage of remote and face-to-face consultations by age group

**Table S1.** Codes to identify remote consultations from observations table

| obs_medcodeid | obs_medterm |
| --- | --- |
| 285327012 | telephone encounter |
| 1780456010 | telephone triage encounter |
| 62141000000119 | telephone call to a patient |
| 600831000000112 | consultation via multimedia |
| 62151000000116 | telephone consultation |
| 11990111000006100 | online triage encounter |
| 11990201000006100 | online triage outcome - video consultation |
| 12991091000006100 | advice given about 2019 novel coronavirus by telephone |
| 13012411000006100 | advice given about sars-cov-2 (severe acute respiratory syndrome coronavirus 2) by telephone |
| 7353351000006110 | telemedicine consultation with patient |
| 8012271000006110 | telephone consultation |
| 12991081000006100 | advice given about wuhan 2019-ncov (novel coronavirus) by telephone |
| 11639701000006100 | asthma nurse specialist telephone encounter |
| 11990141000006100 | online triage outcome - telephone consultation |
| 1778971000006110 | consultation via telemedicine web camera |
| 285332013 | encounter by computer link |
| 7786391000006110 | consultation via video conference |
| 8442071000006110 | econsultation via online application |
| 13012191000006100 | telephone consultation for suspected sars-cov-2 (severe acute respiratory syndrome coronavirus 2) |
| 600851000000117 | consultation via video conference |
| 1849991000006110 | first telephone consultation |
| 63381000000118 | telephone call from a patient |
| 8105181000006110 | consultation by telephone |
| 8344901000006110 | mental health prison inreach team telephone encounter |
| 12990411000006100 | telephone consultation for suspected 2019-ncov (novel coronavirus) |
| 12990421000006100 | telephone consultation for suspected wuhan 2019-ncov (novel coronavirus) |
| 1161431000000110 | telephone consultation for suspected swine flu |
| 8105301000006110 | assessment via video conference |
| 8361131000006110 | consultation via multimedia |
| 8099421000006110 | video-link encounter |
| 12990431000006100 | telephone consultation for suspected 2019 novel coronavirus |
| 12991071000006100 | advice given about 2019-ncov (novel coronavirus) by telephone |
| 11639691000006100 | clinical nurse specialist telephone encounter |
| 1958701000006110 | online communication |
| 1009181000006110 | telephone interpreter used |
| 480941000000110 | nurse telephone triage |
| 1480626017 | telephone follow-up |
| 11990121000006100 | online triage outcome |
| 1995221000006110 | email encounter |
| 1484972014 | patient given telephone advice during surgery hours |
| 1773301000006110 | e-mail consultation |
| 457040019 | patient given telephone advice out of hours |
| 11990211000006100 | online triage outcome - email sent to patient |
| 11990131000006100 | online triage outcome - prescription issued |
| 8105161000006110 | remote consultation |
| 1672891000006110 | telephone call to relative/carer |
| 661661000000111 | e-mail encounter to carer |
| 1672901000006110 | telephone call from relative/carer |
| 1778991000006110 | consultation via sms text message |
| 11990171000006100 | online triage outcome - patient to make appointment online |
| 285396014 | patient 'called' - prevention |
| 2015331000006110 | telehealth monitoring - telephone call to patient |
| 2015351000006110 | telehealth monitoring - telephone call to relative/carer |
| 1161531000000110 | advice given about swine flu by telephone |

**Table S2.**

|  | 2018-19 | 2019-20 | 2020-21 | 2021-22 |
| --- | --- | --- | --- | --- |
| Overall |  |  |  |  |
| 1 - least deprived | 3.88 (3.84-3.91) | 3.93 (3.9-3.97) | 3.63 (3.6-3.66) | 4.28 (4.24-4.32) |
| 2 | 3.93 (3.9-3.97) | 3.97 (3.93-4) | 3.73 (3.69-3.76) | 4.3 (4.26-4.34) |
| 3 | 4.01 (3.97-4.04) | 4.08 (4.05-4.12) | 3.86 (3.83-3.9) | 4.46 (4.42-4.5) |
| 4 | 4.02 (3.99-4.05) | 4.07 (4.03-4.1) | 3.85 (3.82-3.88) | 4.5 (4.46-4.53) |
| 5 - most deprived | 4.24 (4.2-4.27) | 4.18 (4.15-4.22) | 3.93 (3.9-3.96) | 4.46 (4.43-4.5) |
| Face-to-face |  |  |  |  |
| 1 - least deprived | 3.09 (3.06-3.12) | 3.16 (3.14-3.19) | 1.78 (1.76-1.79) | 2.48 (2.46-2.5) |
| 2 | 3.21 (3.18-3.23) | 3.24 (3.21-3.26) | 1.85 (1.84-1.87) | 2.51 (2.48-2.53) |
| 3 | 3.24 (3.21-3.26) | 3.29 (3.26-3.32) | 1.9 (1.88-1.92) | 2.59 (2.57-2.62) |
| 4 | 3.25 (3.23-3.28) | 3.28 (3.26-3.31) | 1.84 (1.82-1.85) | 2.59 (2.56-2.61) |
| 5 - most deprived | 3.46 (3.43-3.48) | 3.41 (3.38-3.44) | 1.8 (1.79-1.82) | 2.54 (2.52-2.56) |
| Remote |  |  |  |  |
| 1 - least deprived | 0.74 (0.73-0.75) | 0.72 (0.71-0.73) | 1.81 (1.8-1.83) | 1.78 (1.76-1.8) |
| 2 | 0.68 (0.67-0.68) | 0.68 (0.67-0.69) | 1.84 (1.82-1.86) | 1.78 (1.76-1.8) |
| 3 | 0.72 (0.71-0.73) | 0.74 (0.73-0.75) | 1.93 (1.91-1.95) | 1.85 (1.83-1.87) |
| 4 | 0.72 (0.71-0.73) | 0.74 (0.74-0.75) | 1.98 (1.96-2) | 1.89 (1.87-1.92) |
| 5 - most deprived | 0.73 (0.72-0.74) | 0.73 (0.72-0.73) | 2.09 (2.07-2.11) | 1.91 (1.89-1.93) |

**Table S3.**

|  | 2018-19 |  | 2019-20 |  | 2020-21 |  | 2021-22 |  |
| --- | --- | --- | --- | --- | --- | --- | --- | --- |
|  | Women | Men | Women | Men | Women | Men | Women | Men |
| Overall |  |  |  |  |  |  |  |  |
| 1 - least deprived | 4.63 (4.58-4.68) | 3.24 (3.21-3.28) | 4.71 (4.66-4.75) | 3.29 (3.25-3.32) | 4.49 (4.45-4.54) | 2.93 (2.9-2.96) | 5.3 (5.24-5.35) | 3.46 (3.42-3.49) |
| 2 | 4.77 (4.72-4.81) | 3.25 (3.21-3.28) | 4.81 (4.76-4.86) | 3.27 (3.23-3.3) | 4.68 (4.63-4.73) | 2.96 (2.93-3) | 5.4 (5.35-5.46) | 3.43 (3.39-3.46) |
| 3 | 4.87 (4.82-4.92) | 3.3 (3.26-3.33) | 4.97 (4.92-5.02) | 3.36 (3.32-3.39) | 4.87 (4.82-4.92) | 3.07 (3.04-3.1) | 5.62 (5.56-5.67) | 3.54 (3.51-3.58) |
| 4 | 4.91 (4.86-4.96) | 3.29 (3.26-3.32) | 4.98 (4.93-5.03) | 3.32 (3.29-3.36) | 4.88 (4.83-4.93) | 3.04 (3.01-3.07) | 5.69 (5.64-5.75) | 3.55 (3.51-3.58) |
| 5 - most deprived | 5.24 (5.19-5.29) | 3.42 (3.39-3.46) | 5.18 (5.13-5.23) | 3.38 (3.34-3.41) | 5.04 (4.99-5.09) | 3.06 (3.03-3.09) | 5.72 (5.66-5.78) | 3.48 (3.45-3.52) |
| Face-to-face |  |  |  |  |  |  |  |  |
| 1 - least deprived | 3.67 (3.63-3.71) | 2.6 (2.57-2.63) | 3.76 (3.72-3.8) | 2.66 (2.63-2.69) | 2.17 (2.15-2.2) | 1.45 (1.44-1.47) | 3.04 (3.01-3.08) | 2.02 (2-2.04) |
| 2 | 3.87 (3.83-3.9) | 2.66 (2.63-2.69) | 3.9 (3.86-3.94) | 2.68 (2.65-2.71) | 2.3 (2.28-2.32) | 1.49 (1.48-1.51) | 3.12 (3.09-3.16) | 2.01 (1.99-2.03) |
| 3 | 3.91 (3.87-3.95) | 2.67 (2.65-2.7) | 3.98 (3.94-4.02) | 2.72 (2.69-2.75) | 2.37 (2.34-2.39) | 1.53 (1.51-1.54) | 3.24 (3.21-3.28) | 2.08 (2.05-2.1) |
| 4 | 3.95 (3.91-3.99) | 2.68 (2.65-2.7) | 3.99 (3.95-4.03) | 2.7 (2.67-2.73) | 2.3 (2.27-2.32) | 1.47 (1.45-1.48) | 3.25 (3.21-3.28) | 2.06 (2.04-2.08) |
| 5 - most deprived | 4.25 (4.21-4.29) | 2.81 (2.78-2.84) | 4.2 (4.16-4.24) | 2.77 (2.74-2.8) | 2.29 (2.26-2.31) | 1.42 (1.41-1.44) | 3.23 (3.2-3.27) | 2 (1.98-2.02) |
| Remote |  |  |  |  |  |  |  |  |
| 1 - least deprived | 0.92 (0.9-0.93) | 0.6 (0.59-0.6) | 0.9 (0.89-0.91) | 0.58 (0.57-0.59) | 2.28 (2.25-2.3) | 1.45 (1.43-1.47) | 2.23 (2.2-2.26) | 1.42 (1.4-1.44) |
| 2 | 0.85 (0.84-0.86) | 0.54 (0.53-0.54) | 0.86 (0.85-0.88) | 0.54 (0.53-0.55) | 2.34 (2.32-2.37) | 1.44 (1.43-1.46) | 2.26 (2.23-2.29) | 1.4 (1.38-1.42) |
| 3 | 0.91 (0.9-0.93) | 0.57 (0.56-0.58) | 0.94 (0.93-0.95) | 0.58 (0.58-0.59) | 2.46 (2.43-2.49) | 1.51 (1.49-1.53) | 2.36 (2.33-2.39) | 1.45 (1.43-1.47) |
| 4 | 0.92 (0.9-0.93) | 0.56 (0.55-0.57) | 0.96 (0.94-0.97) | 0.58 (0.57-0.59) | 2.56 (2.53-2.59) | 1.54 (1.52-1.56) | 2.44 (2.41-2.47) | 1.47 (1.45-1.49) |
| 5 - most deprived | 0.94 (0.93-0.95) | 0.57 (0.56-0.57) | 0.94 (0.93-0.95) | 0.56 (0.55-0.57) | 2.73 (2.69-2.76) | 1.61 (1.59-1.63) | 2.49 (2.46-2.52) | 1.47 (1.45-1.49) |

## References

1. Bakhai M, Croney L, Waller O, Henshall N FC. Using Online Consultations In Primary Care Implementation Toolkit. 2020.

2. Marshall M, Howe A, Howsam G, Mulholland M, Leach J. COVID-19: A danger and an opportunity for the future of general practice. Vol. 70, British Journal of General Practice. 2020.

3. Murphy M, Scott LJ, Salisbury C, Turner A, Scott A, Denholm R, et al. Implementation of remote consulting in UK primary care following the COVID-19 pandemic: A mixed-methods longitudinal study. British Journal of General Practice. 2021 Mar 1;71(704):E166–77.

4. Velasquez D, Mehrotra A. Ensuring the growth of telehealth during COVID-19 does not exacerbate disparities in care. Health Affairs Blog. 2020;1–8.

5. Nussbaum C, Massou E, Fisher R, Morciano M, Harmer R, Ford J. Inequalities in the distribution of the general practice workforce in England: A practice-level longitudinal analysis. BJGP Open. 2021 Oct 1;5(5):1–10.

6. Ruth P, Emma F, Charlotte P, James M, David B, John F. Inequalities in General Practice Remote Consultations: A Systematic Review. BJGP Open. 2021 Mar 12;12(12):1–12.

7. Howells K, Amp M, Burrows M, Brown J, Brennan R, Dickinson J, et al. Remote primary care during the COVID-19 pandemic for people experiencing homelessness: a qualitative study. British Journal of General Practice. 2022 Apr 4;BJGP.2021.0596.

8. Joy M, McGagh D, Jones N, Liyanage H, Sherlock J, Parimalanathan V, et al. Reorganisation of primary care for older adults during COVID-19: A cross-sectional database study in the UK. British Journal of General Practice. 2020 Aug 1;70(697):E540–7.

9. Clinical Practice Research Datalink. CPRD Aurum May 2022 (Version 2022.05.001) [Internet]. 2022 [cited 2022 Nov 15]. Available from: https://cprd.com/cprd-aurum-may-2022-dataset

10. Watt T, Sullivan R, Aggarwal A. Primary care and cancer: an analysis of the impact and inequalities of the COVID-19 pandemic on patient pathways. BMJ Open. 2022 Mar 1;12(3):e059374.

11. Hobbs FDR, Bankhead C, Mukhtar T, Stevens S, Perera-Salazar R, Holt T, et al. Clinical workload in UK primary care: a retrospective analysis of 100 million consultations in England, 2007–14. The Lancet [Internet]. 2016 Jun 4 [cited 2022 May 4];387(10035):2323–30. Available from: http://dx.doi.org/10.1016/

12. Green MA, McKee M, Katikireddi SV. Remote general practitioner consultations during COVID-19. Vol. 4, The Lancet Digital Health. Elsevier Ltd; 2022. p. e7.

13. Foley KA, Maile EJ, Bottle A, Neale FK, Viner R, Kenny SE, et al. Impact of covid-19 on primary care contacts with children and young people aged 0-24 years in England; longitudinal trends study 2015-2020. British Journal of General Practice. 2022 Apr 4;BJGP.2021.0643.

14. Ledger J, Bakhai M. The Temporal Dimensions of Health Technology Adoption During the Covid-19 Pandemic: Revisiting Roger’s Diffusionist Innovation Theory BT - Organising Care in a Time of Covid-19: Implications for Leadership, Governance and Policy. In: Waring J, Denis JL, Reff Pedersen A, Tenbensel T, editors. Cham: Springer International Publishing; 2021. p. 245–73. Available from: https://doi.org/10.1007/978-3-030-82696-3_12

15. Baines R, Tredinnick-Rowe J, Jones R, Chatterjee A. Barriers and Enablers in Implementing Electronic Consultations in Primary Care: Scoping Review. J Med Internet Res [Internet]. 2020 Nov 12 [cited 2022 Nov 2];22(11):e19375. Available from: https://www.jmir.org/2020/11/e19375

16. Rezel-Potts E, L’Esperance V, Gullifiord M. Antimicrobial stewardship during the COVID-19 epidemic: population-based cohort study and interrupted time series analysis in the United Kingdom. British Journal of General Practice [Internet]. 2021 [cited 2021 Apr 11];71(706):E331–8. Available from: https://doi.org/10.3399/BJGP.2020.1051

17. Cookson R, Propper C, Asaria M, Raine R. Socio-Economic Inequalities in Health Care in England. Fisc Stud. 2016 Sep 1;37(3–4):371–403.

18. Fisher R, Dunn P, Gershlick B, Asaria M, Thorlby R. Level or not? 2020 Sep 26 [cited 2022 Nov 15]; Available from: https://www.health.org.uk/publications/reports/level-or-not

19. Nussbaum C, Massou E, Fisher R, Morciano M, Harmer R, Ford J. Inequalities in the distribution of the general practice workforce in England: a practice-level longitudinal analysis. BJGP Open. 2021 Oct;5(5):1–10.

20. Phiri P, Delanerolle G, Al-Sudani A, Rathod S. COVID-19 and black, Asian, and minority ethnic communities: A complex relationship without just cause. JMIR Public Health Surveill. 2021;7(2).

21. Simpson RM, Knowles E, O’Cathain A. Health literacy levels of British adults: a cross-sectional survey using two domains of the Health Literacy Questionnaire (HLQ). BMC Public Health. 2020;20(1).

22. Francis-Devine B. Poverty in the UK: statistics [Internet]. 2022 [cited 2022 Nov 15]. Available from: https://researchbriefings.files.parliament.uk/documents/SN07096/SN07096.pdf

23. The Health Foundation, Ipsos-MORI. Public perceptions of health and social care (November–December 2021) - The Health Foundation [Internet]. [cited 2022 Oct 27]. Available from: https://www.health.org.uk/publications/public-perceptions-of-health-and-social-care-november-december-2021

24. Davies AR, Honeyman M, Gann B. Addressing the digital inverse care law in the time of COVID-19: Potential for digital technology to exacerbate or mitigate health inequalities. J Med Internet Res [Internet]. 2021 Apr 7 [cited 2021 Oct 21];23(4):e21726. Available from: https://www.jmir.org/2021/4/e21726

25. Morales DR, Ali SN. COVID-19 and disparities affecting ethnic minorities. The Lancet. 2021 May 8;397(10286):1684–5.

26. Head A, Fleming K, Kypridemos C, Schofield P, Pearson-Stuttard J, O’Flaherty M. Inequalities in incident and prevalent multimorbidity in England, 2004–19: a population-based, descriptive study. Lancet Healthy Longev. 2021;2(8).

27. Rosen R, Wieringa S, Greenhalgh T, Leone C, Rybczynska-Bunt S, Hughes G, et al. Clinical risk in remote consultations in general practice: findings from in-COVID-19 pandemic qualitative research. BJGP Open [Internet]. 2022 Sep 1 [cited 2022 Oct 17];6(3):BJGPO.2021.0204. Available from: https://bjgpopen.org/content/6/3/BJGPO.2021.0204

28. Wang Y, Hunt K, Nazareth I, Freemantle N, Petersen I. Do men consult less than women? An analysis of routinely collected UK general practice data. BMJ Open. 2013;3(8).

29. Watt T, Raymond A, Rachet-Jacquet L. Quantifying health inequalities in England. London, England; 2022.

30. Darley S, Coulson T, Peek N, Moschogianis S, Veer SN van der, Wong DC, et al. Understanding How the Design and Implementation of Online Consultations Affect Primary Care Quality: Systematic Review of Evidence With Recommendations for Designers, Providers, and Researchers. J Med Internet Res 2022;24(10):e37436 https://www.jmir.org/2022/10/e37436 [Internet]. 2022 Oct 24 [cited 2022 Nov 16];24(10):e37436. Available from: https://www.jmir.org/2022/10/e37436

